# Colorimetric and fluorometric reverse-transcription loop-mediated isothermal amplification (RT-LAMP) assay for diagnosis of SARS-COV-2

**DOI:** 10.1101/2022.08.30.22279408

**Authors:** Galyah Alhamid, Huseyin Tombuloglu, Dalal Motabagani, Dana Motabagani, Ali A. Rabaan, Kubra Unver, Gabriel Dorado, Ebtesam Al-Suhaimi, Turgay Unver

**Affiliations:** Department of Genetics Research, Institute for Research and Medical Consultations (IRMC), Imam Abdulrahman Bin Faisal University, 31441, Dammam, Saudi Arabia; College of Medicine, King Faisal University, 31982, Al-Ahsa, Saudi Arabia; Molecular Diagnostic Laboratory, Johns Hopkins Aramco Healthcare, Dhahran, Saudi Arabia; College of Medicine, Alfaisal University, Riyadh 11533, Saudi Arabia; Department of Public Health and Nutrition, the University of Haripur, Haripur 22610, Pakistan; Ficus Biotechnology, Ostim OSB Mah, 100. Yil Blv, No: 55, Yenimahalle, Ankara, Turkey; Dep. Bioquímica y Biología Molecular, Campus Rabanales C6-1-E17, Campus de Excelencia Internacional Agroalimentario (ceiA3), Universidad de Córdoba, 14071 Córdoba, Spain; Biology Department, College of Science and Institute of Research and Medical Consultations (IRMC), Imam Abdulrahman Bin Faisal University, 31441, Dammam, Saudi Arabia

**Keywords:** SARS-CoV-2, RT-LAMP, coronavirus, colorimetric, fluorometric, diagnosis, COVID-19

## Abstract

The coronavirus disease 2019 (COVID-19) caused by the Severe Acute Respiratory Syndrome Coronavirus-2 (SARS-CoV-2) has caused millions of infections and deaths worldwide, since it infected humans almost three years ago. Improvements of current assays and development of new rapid tests or to diagnose SARS-CoV-2 are urgent. Reverse-transcription loop-mediated isothermal amplification (RT-LAMP) is a rapid and propitious assay, allowing to detect both colorimetric and/or fluorometric nucleic-acid amplifications. This study describes the analytical and clinical evaluation of RT-LAMP assay for detection of SARS-CoV-2, by designing LAMP primers targeting *N* (nucleocapsid phosphoprotein), *RdRp* (polyprotein), *S* (surface glycoprotein) and *E* (envelope protein) genes. The assay’s performance was compared with the gold-standard RT-PCR, yielding 94.6% sensitivity and 92.9% specificity. Among the tested primer sets, the ones for *S* and *N* genes had the highest analytical sensitivity, showing results in about 20 minutes. The colorimetric and fluorometric comparisons revealed that the latter is faster than the former. The limit of detection (LoD) of RT-LAMP reaction in both assays is 50 copies/µl of reaction mixture. However, the simple eye-observation advantage of the colorimetric assay (with a color change from yellow to red) serves a promising on-site point-of-care testing method anywhere, including, for instance, laboratory and in-house applications.

## Introduction

As of late 2019, the outbreak of a novel human virus, <u>S</u>evere <u>A</u>cute <u>R</u>espiratory <u>S</u>yndrome <u>Co</u>rona<u>v</u>irus <u>2</u> (SARS-CoV-2), which causes the <u>co</u>rona<u>vi</u>rus <u>d</u>isease 20<u>19</u> (COVID-19), has infected more than 562 million people all around the world, at the time of writing, as shown by the Coronavirus Resource Center at the Johns Hopkins University School of Medicine <https://coronavirus.jhu.edu/map.html>. Death numbers have reached over six million as of July 2022 (*COVID Live - Coronavirus Statistics - Worldometer* 2022). The development of detection kits and tools has been extremely important for early diagnosis of such disease. Besides *in vitro* molecular tests, current coronavirus diagnosis protocols are based on clinical symptom. Yet, the latter resembles the ones of influenza and common cold, pointing to the need of accurate and rapid molecular tests for reliable diagnosis.

Among currently available *in vitro* diagnostics (IVD), polymerase chain-reaction (PCR)-based kits, serological tests and metagenomic next-generation sequencing (mNGS) were the validated methods to detect SARS-CoV-2. Serological tests detect SARS-CoV-2 antigen or antibodies [Immunoglobulins G and M (IgG and IgM)] from patients’ sera to identify recent exposure or immunity (Behera et al. 2021). One of the most popular serological tests to detect SARS-CoV-2 is the enzyme-linked immunosorbent assay (ELISA) (Iruretagoyena et al. 2021; Vernet et al. 2021). It relies on binding antibodies from patient’s sera or blood to antigens coated on microtiter wells, resulting in complexes that changes the color of the solution after adding a substrate (Aileni et al. 2022). Also, lateral-flow assays caught the attention of researchers, because they are small portable devices that can be used as a point-of-care (POC) testing technique. These assays use the immunochromatography principle. Thus, drops of a patient sample (blood, swabs, or saliva) flow through a test pad conjugated with IgG and/or IgM, which form antigen-antibody complexes if the person is infected (Grant et al. 2020; Li et al. 2020). In addition, neutralization assays measure the ability of cultured cells or patients’ samples of producing neutralizing antibodies, when exposed to SARS-CoV-2 antigens (Abe et al. 2020). Aside from serological tests, clustered regularly-interspaced short-palindromic repeats (CRISPR)-based technology along with its associated enzymes (Cas 9, Cas 12, and Cas13) have been widely invested, showing high accuracy in COVID-19 diagnosis (Broughton et al. 2020; F. Zhang et al. 2020). Biosensors have also caught attention during the COVID-19 pandemic. These devices include a transduction element that quantifies biochemical reactions from antigen-antibody complexes. They generate an output signal from bioreceptor element, including immobilized antibodies or nucleic acids, being analyzed by a detection system (Aileni et al. 2022; Behera et al. 2021). Biosensors effectively demonstrated high sensitivity and specificity against SARS-CoV-2, in addition to providing real-time detection (Chaibun et al. 2021; Seo et al. 2020).

PCR-based tests are useful for initial diagnosis. Yet, they require expert personnel and equipped laboratories that can substantially delay testing time even for days, depending on laboratory turnaround and sample-shipping time (Sheridan 2020). On the other hand, serological tests could rapidly diagnose within minutes, but they can only identify persons with detectable blood antibody levels, and they take a longer time to develop (Ravi et al. 2020). Others, like CRISPR-based techniques are prone to false results when viral mutations are recurrent. Finally, biosensors require a considerably higher amount of analyte to result in a detectable response (Abid et al. 2021). Therefore, due to the limitations of the currently available methodologies, development of new cost- and time-efficient approaches with on-site testing capability is desirable. In this regard, loop-mediated isothermal amplification (LAMP)-based diagnostics are having considerable attention, thanks to their simple design, easy implementation, and POC testing capability, as well as reduced cost and testing time to generate results (Wong et al. 2018).

LAMP is a nucleic-acid amplification technology using four to six specially-designed primers —forward outer (F3), forward inner (FIP), loop forward (LF), backward outer (B3), backward inner (BIP) and loop backward (LB)— and a DNA polymerase with high strand-displacement activity, at 60-65 ^º^C constant temperature (Kashir and Yaginuddin 2020). It can also be combined with reverse transcription to target RNA (RT-LAMP), where reverse transcription and amplification take place simultaneously (Yu et al. 2020). LAMP results can be detected by various approaches, such as change in turbidity caused by magnesium pyrophosphate precipitate (Mori et al. 2004), probe-based with nanoparticles (Bhat et al. 2019), strip pH indicators (Bhat and Rao 2020), fluorescence reading by using intercalating dyes (Dao Thi et al. 2020), gel electrophoresis followed by UV detection (Aoki et al. 2021) or even with the naked-eye, as a reaction-color change (Zhang et al. 2020).

*In-situ* testing capability of RT-LAMP, even with equivalent performance to commercial PCR-based tests, broadened the convenience and scope of its applicability. Successful applications of LAMP, coupled with different methodologies, have been demonstrated in many studies, including COVID-19 (Yu et al. 2020; Zhang et al. 2020; Dao Thi et al. 2020; El-Tholoth et al. 2020; Aoki et al. 2021), MERS-CoV (Shirato et al. 2018), EcoV (Nemoto et al. 2015), SADS-CoV (Wang et al. 2018), FcoV (Stranieri et al. 2017), HIV-1 (Rudolph et al. 2015), and many others.

Among the current coronavirus diagnostic technologies such as RT-PCR, LAMP-based ones are highly prioritized, due to their lower cost and time efficiency, DNA/RNA targeting ability, simple design and easy implementation. Also, such latter assays do not require expensive qPCR instrumentation when used as colorimetric. The current study demonstrates both colorimetric and fluorometric LAMP assays by targeting *N, RdRp, S* and *E* genes of SARS-CoV-2 genomes, belonging to the most recent variants of concern (VOC), including B.1.1.7 (Alpha), B.1.351 (Beta), P.1 (Gamma), B.1.617.2 (Delta), and B.1.1.529 (Omicron).

## Materials and methods

### RT-LAMP primer design

A total of 1,000 SARS-CoV-2 genomes belonging to VOC, including the five described in the previous paragraph were downloaded from Global Initiative on Sharing Avian Influenza Data (GISAID) <https://www.gisaid.org>. Virus genome sequences were selected from different countries, belonging to five continents. In addition, genome sequences of SARS-CoV relatives such as AY278491.2, AY502924.1, AY502927.1 AY559094.1, AY613947.1 and NC_004718.3 were downloaded from the National Center for Biotechnology Information (NCBI) database <https://www.ncbi.nlm.nih.gov/sars-cov-2>. Sequences were first aligned by using MAFFT software version 7?? <https://mafft.cbrc.jp/alignment/server> (Katoh et al. 2019). Then, mutation sites (polymorphisms) on the genes were screened by using JalView program version 2.11.1.3 <https://www.jalview.org> (Waterhouse et al. 2009). Then, 100% conserved (non-mutated; monomorphic) regions specific to SARS-CoV-2, but non-homolog for other viruses, were selected as potential primer-binding sites. PrimerExplorer program version 5 <https://primerexplorer.jp/e> was used to design six LAMP primers in each set, targeting conserved region of specific genes (*RdRp, N, E* and *S*, genes; Table S1). Primers designed for the *S* gene have also been used by others (Yan et al. 2020). Since the target region is narrow and the same design program was used, it was expected to have the same primer design. To avoid forming secondary structures (homodimers, hetero-dimers and hairpins), OligoAnalyzer tool from Integrated DNA Technologies (IDT) <https://eu.idtdna.com/pages/tools/oligoanalyzer> was used. Many different primer sets were designed and tested. Primers showing the best performance without primer dimers in no-template control (NTC) were selected. Primers were synthesized by Alpha DNA (Montreal, Canada) <http://www.alphadna.com>, desalted and lyophilized. Concentrations of oligonucleotides in the reaction mixtures (20 µl) were adjusted as 0.2 (2 µM), 0.2 (2 µM), 1.6 (16 µM), 1.6 (16 µM), 0.8 (8 µM) and 0.8 (8 µM), for F3, B3, FIP, BIP, LB and LF primers, respectively.

### Samples and RNA extraction

This study was approved by the Institutional Review Board (IRB) at Imam Abdulrahman bin Faisal University (IAU) with IRB no. IRB-2020-13-406. According to the institutional ethics committee regulations, no participants’ consent was required, since samples used in this study were post-diagnostic leftover tests, being de-identified. The nasopharyngeal/throat (NP/T) swabs of COVID-19-suspected patients were collected and transferred into a liquid Viral Transport Medium (VTM) from Copan (Murrieta, CA, USA) <https://www.copanusa.com>. Specimens were kept at 4 DC for less than eight hours before RNA extraction. RNA isolation was carried out according to the suggested protocol of QIAamp Viral RNA mini kit from Qiagen (Hilden, Germany) <https://www.qiagen.com>. The SARS-CoV-2 Omicron synthetic-RNA control (Twist synthetic RNA control 51; EPI_ISL_7718520) was obtained from Twist Bioscience (South San Francisco, CA, USA) <https://www.twistbioscience.com>. The specificity of the primers was determined by testing other respiratory viruses’ RNA extracted from clinical samples, including parainfluenza virus 3, enterovirus, rhinovirus, human metapneumovirus A+B, parainfluenza virus 4, bocavirus and coronavirus 229 E.

### RT-LAMP assay

RT-LAMP reaction mixtures were composed of the following components: 10x LavaLAMP RNA buffer and 1 µl LavaLAMP RNA enzyme from Lucigen (Middleton, WI, USA) <https://www.lucigen.com>, 100 mM MgSO_4_, 10 mM dNTP, 2.5 µl primer mixture, 5 µl template and ddH_2_O up to 20 µl. For colorimetric assays, 2.5 mM Neutral Red was added to the mixture, being replaced with Green Fluorescent dye from Lucigen in the fluorometric assays. Colorimetric RT-LAMP mixtures were incubated in a thermal block at 70 ^º^C for 45 minutes. Reactions were terminated at 95 ^º^C for 5 minutes, and immediately cooled down at 4 ^º^C, being crucial to allow post-reaction analyses and validations. Fluorometric RT-LAMP was performed in a 7500 Fast Real-Time PCR System from Thermo Fisher Scientific (Waltham, MA, USA) <https://www.thermofisher.com>, using 80 cycles of 70 °C for 30 seconds. Fluorescence readings were performed at amplification steps of each cycle. FAM filter was selected as the reporter dye channel. In addition, assays were tested by using different light cyclers, such as StepOnePlus Real-Time PCR System from the same manufacturer, CFX96 Touch Real-Time PCR Detection System from Bio-Rad Laboratories (Hercules, CA, USA) <https://www.bio-rad.com>, Q2000 Real-Time PCR System from LongGeneScientific Instruments (Hangzhou, China) <http://en.longgene.com> and LightCycler 480 System from Hoffmann-La Roche (Basel, Switzerland) <https://diagnostics.roche.com>.

### Validation of results by RT-PCR

The RT-PCR was applied according to our previous protocol (Tombuloglu et al. 2021, 2022). In multiplex RT-PCR reactions, viral *RdRp* and *N*, and human *RP* genes were targeted. A positive control containing synthetic copies of each target gene was used (EURM-019, single stranded SARS–CoV-2 RNA fragments) from the European Commission, Joint Research Centre Directorate F – Health, Consumers and Reference Materials (Geel, Belgium) <https://crm.jrc.ec.europa.eu/p/EURM-019>. For NTC, RNase/DNase-free ddH_2_O was added instead of the RNA template. To validate LAMP results, reaction mixtures were segregated using 2% agarose gel electrophoresis (AGE), stained with VisualaNA (A) DNA Stain from Molequle-On (Auckland, New Zealand) <http://molequle-on.com>. Gels were imaged under a UV-trans illuminator (ChemiDoc XRS+ System with Image Lab Software) from Bio-Rad Laboratories.

## Results

### Colorimetric and fluorometric identification of positive samples

Fig. 1 depicts tested individuals, according to the RT-LAMP assay. Different primer sets targeting *N, S, RdRp* and *E*, genes were evaluated, via both colorimetric and fluorometric identification. Using the pH-sensitive neutral red dye, a color change to red was evident in positive control (PC) samples after colorimetric RT-LAMP reactions, while NTC was yellow. Also, in the fluorometric identification, PC samples produced a fluorescence amplification after ∼40 cycles (around 20 minutes), and no amplification was evident in NTC of all tested primer sets. Development of yellow to red color was more pronounced in positive reaction tubes prepared with *N* or *S* primer sets, compared to *RdRp* and *E* ones (Fig. 1a-d). However, there was no significant difference in Ct scores of positive amplification curves among tested target genes (Fig. 1a1-d1).

**Fig. 1.**
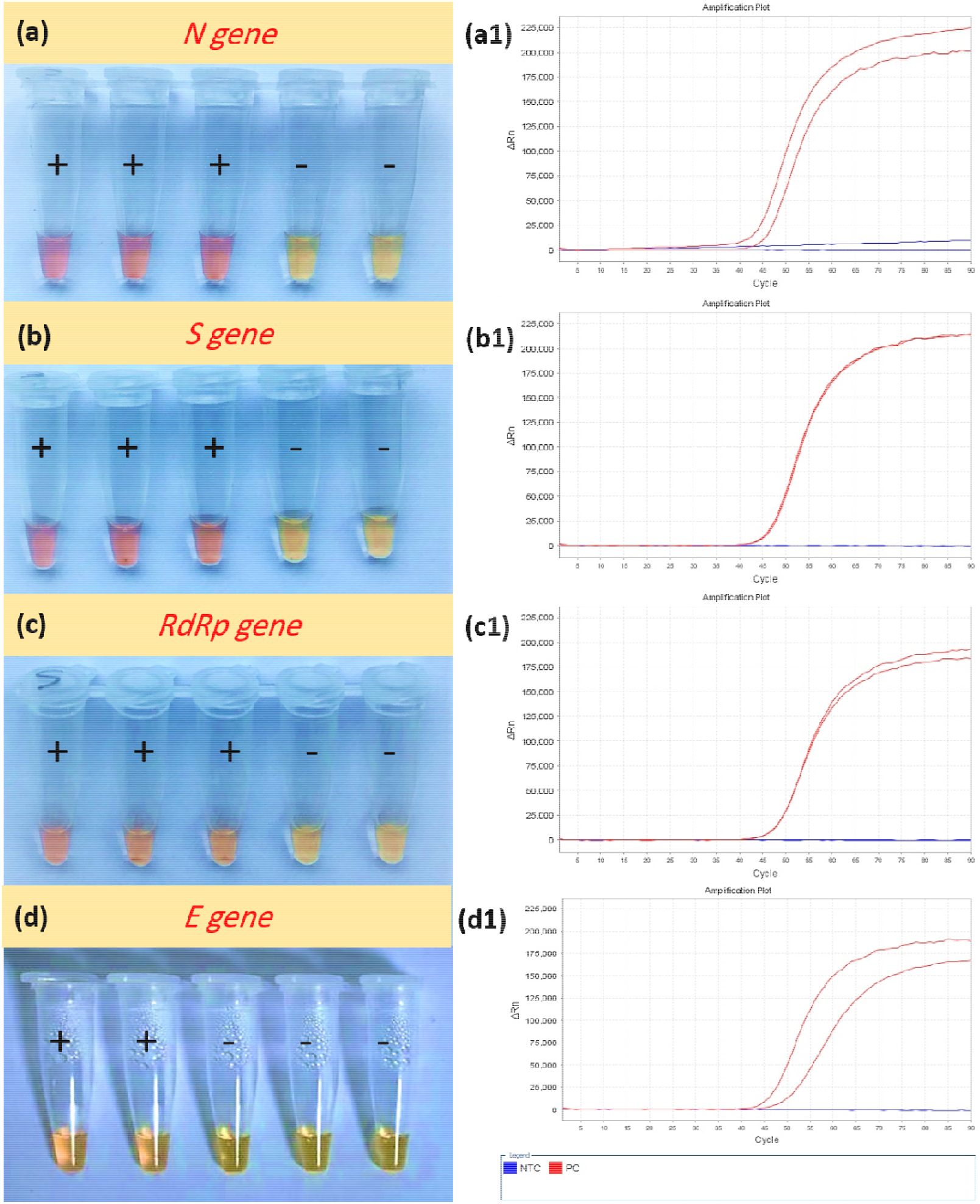
RT-LAMP based **(a-d)** colorimetric and **(a1-d1)** fluorometric identification of SARS-CoV-2 positive controls (PC, +) and negative template controls (NTC, –) samples, by using *N* **(a, a1)**, *S* **(b, b1)**, *RdRp* **(c, c1)** and *E* **(d, d1)** gene primer sets. SARS-CoV-2 positive samples undergo color change from yellow to red/pink in the colorimetric-based reactions, producing amplification curves in fluorometric-identification methods.

### RT-LAMP assay on clinical samples

Since colorimetric identification leads to more noticeable color development for *N* and *S* primer sets, performance of those primer sets was tested on clinical specimens (n = 51), using both colorimetric and fluorometric RT-LAMP assays. Results were then validated using AGE, after colorimetric RT-LAMP reactions. Fig. 2 demonstrates colorimetrically- and fluorometrically-tested specimens (S1-S51), highlighting color differences between positives and negatives, comparing to PC and NTC in each sample set. Among the 51 tested samples, RT-LAMP assays agreed with RT-qPCR results on 48 samples (94.1%), as 34 are detected as SARS-CoV-2 positives and 14 negatives. Three samples were found inconclusive. Thus, S18 was positive in colorimetric RT-LAMP, but negative in RT-qPCR. On the other hand, S30 and S32 were negative in colorimetric RT-LAMP, but positive in RT-qPCR. Therefore, S18 must be regarded as positive, since amplicon formation was evident via color change, being undetected by RT-qPCR (Fig. S1). Color intensities varied among these samples, based on the amount of viral RNA present (Fig. 2a). Also, some of these samples were left at room temperature to check if any further color was developed, as some of them had an immediate color change (within 20 minutes), whereas others took over 60 minutes, thus causing some variety in color intensity. Nonetheless, results were in agreement with fluorometric identification (94.1%), where positive specimens produced fluorescence amplifications with high fluorescence intensities (Fig. 2b). Ct values in positive specimens varied between 35 and 75, and no Ct value was evident in NTC (Fig. 2c). It should be taken into consideration that differences in Ct values may also be due to different viral loads among specimens.

**Fig. 2.**
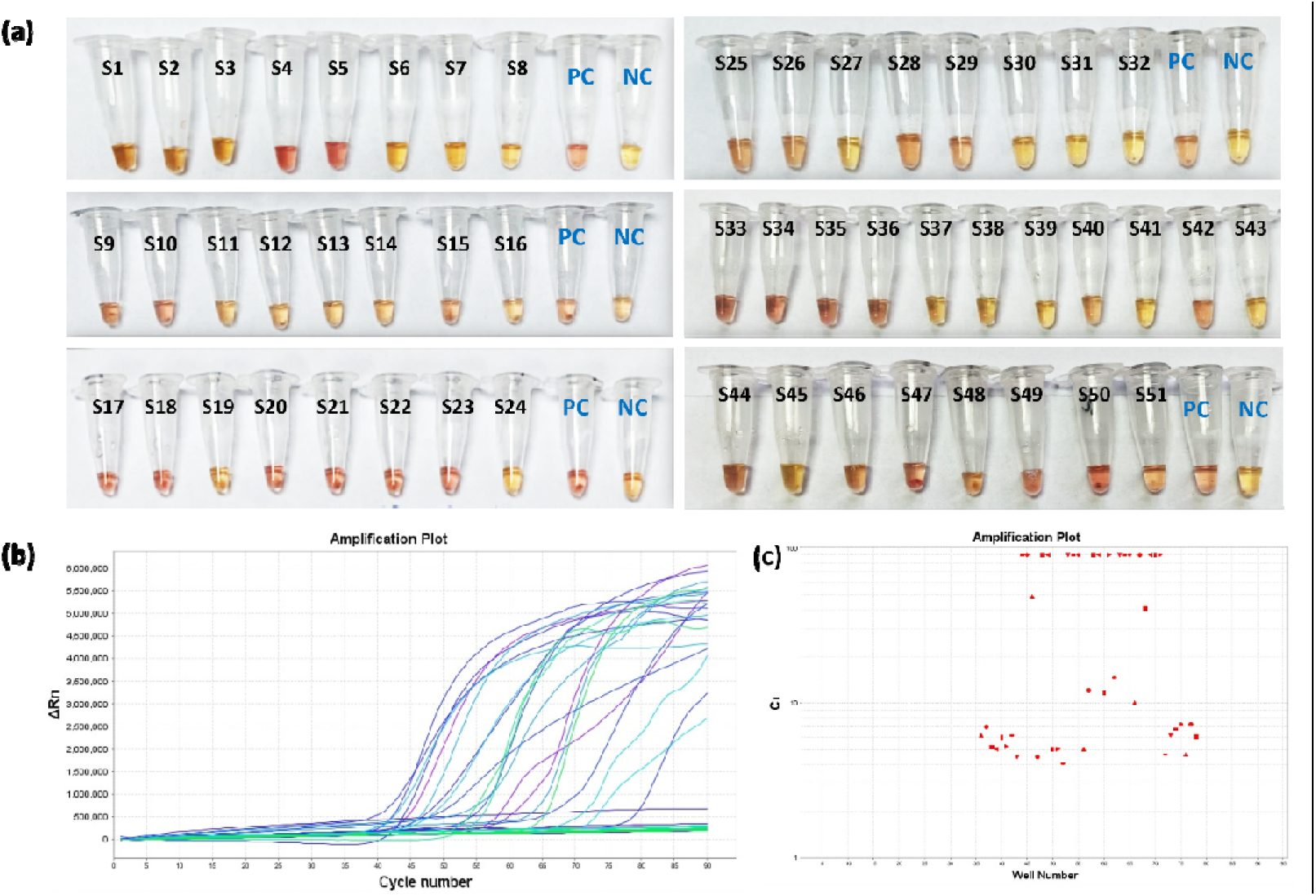
**(a)** Colorimetric and **(b)** fluorometric *S*-gene-specific RT-LAMP assays applied on SARS-CoV-2-positive and -negative RNA specimens. Positive control (PC), negative control (NC). **(c)** Ct spectrum of samples.

In order to confirm both colorimetric and fluorometric results, colorimetric RT-LAMP products were loaded in a 2% agarose gel and visualized under UV trans-illuminator. Amplification products with different sizes appeared for SARS-CoV-2 positive and PC specimens. No bands were evident in negative samples and NTC, confirming successful amplification of SARS-CoV-2 *S* gene in the former. Fig. 3 shows agreement between colorimetric, fluorometric and gel electrophoresis techniques on the patients’ specimens, along with PC and NTC.

**Fig. 3.**
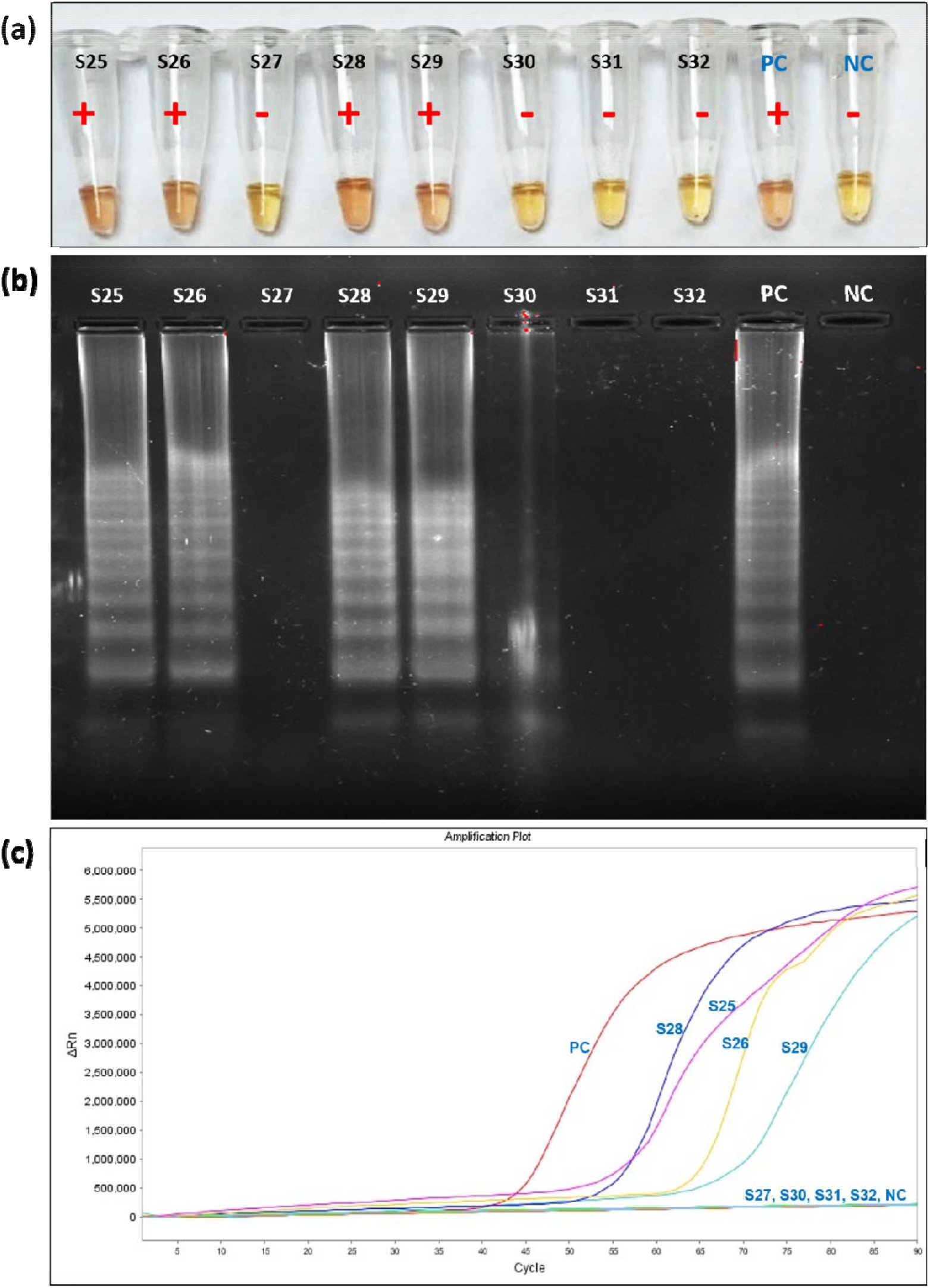
Triple confirmation of *S*-specific LAMP primer sets. **(a)** Colorimetric **(b)** agarose gel electrophoresis and **(c)** fluorometric, showing agreement among these techniques.

### Assay sensitivity and specificity of primer sets

SARS-CoV-2 RNA with known concentration (EURM-019, single stranded SARS– CoV-2 RNA fragments) were serially diluted (1, 10^1^, 10^2^, 10^3^, 10^4^, 10^5^ and 10^6^ fold) to determine the limit of detection (LOD) of RT-LAMP reactions. Fig. 4 shows the LOD of colorimetric and fluorometric RT-LAMP assays, targeting the *S* gene. Results showed that color development from yellow to red is prominent in 1, 10^1^, 10^2^ and 10^3^ fold dilutions. The limit of detection corresponds to 50 copies/µl in reactions, for the 10^3^-fold diluted specimen (Fig. 4a). As expected, samples with higher viral load produced more prominent or darker red color, as compared to lower ones. Likewise, Ct values in fluorometric identifications progressively increased with an increasing dilution factor, further confirming colorimetric results (Fig. 4b). Besides, the color intensity and Ct values were highly dependent on viral load. Once reactions were carried out, they were subjected to AGE in 2% agarose and visualized under a UV trans-illuminator, to further check if band-pattern intensities decreased with reduced viral loads (Fig. 4c). Results indicate a ladder-type banding pattern in detected dilutions, while no bands appeared for others. In summary, there was agreement between agarose gel electrophoresis, fluorometric and colorimetric identifications.

**Fig. 4.**
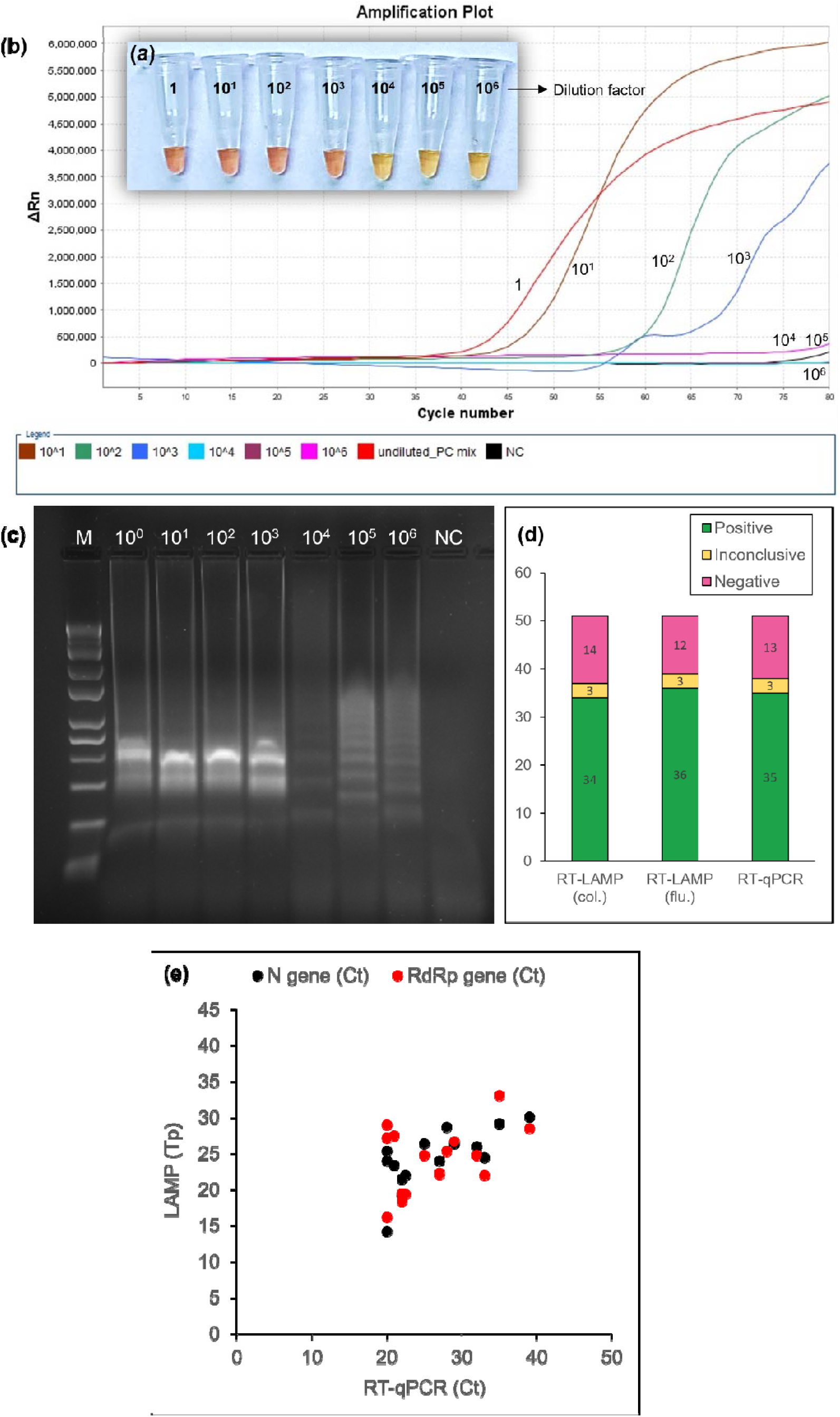
Assay sensitivity of **(a)** colorimetric and **(b)** fluorometric RT-LAMP tests, targeting *S* gene. Dilution factors are represented on the graph. **(c)** Agarose gel electrophoresis of diluted samples (from 10^0^ to 10^6^ fold). M and NC denote marker and negative control, respectively. **(d)** Comparison of colorimetric (col.) and fluorometric (flu.) RT-LAMP with RT-qPCR assays. Among 51 tested samples, RT-LAMP assays, both colorimetric and fluorometric, agreed with RT-qPCR on 48 samples, as 34 positives and 14 negatives. Three samples were found inconclusive. **(e)** Comparison of time to positive (T_p_) of RT-LAMP and cycle threshold (Ct) of RT-qPCR assays, based on *N* (black dots) and *RdRp* (red dots) genes.

When compared with RT-PCR, RT-LAMP assays result in 94% agreement (48 over 51 tested samples) with both fluorometric and colorimetric identifications, where three samples were found inconclusive (Fig. 4d). For instance, sample 18 (S18) was not detected as positive in multiplex RT-qPCR simultaneously targeting both *N* and *RdRp* genes, while *RP* amplification was shown. This reflects an amplification pattern of a negative specimen (Fig. S1). However, both fluorometric and colorimetric RT-LAMP reactions generated positive results, which conflicts with the ones of RT-qPCR. Opposite patterns were found for S30 and S32 specimens (Fig. S1), being therefore considered as inconclusive. As a result, RT-LAMP colorimetric assays showed sensitivity of 94.6%, specificity of 92.9% and accuracy of 94.1%. Among the tested 51 clinical specimens, the assay’s positive predictive value (PPV) was 97.2% and the negative predictive value was 86.7% (Table S2; S3). In addition, RT-LAMP time-to-positive (T_p_) was compared with the Ct values of *N* and *RdRp* genes by RT-qPCR (Fig. 4e) (14 positives and 16 negative clinical samples). Fluorometric LAMP showed 100% agreement with RT-qPCR results.

Similarly, the *N* gene primer-set was tested against SARS-CoV-2 positive (PC) and negative (NC) specimens, using the fluorescence RT-LAMP detection technique. This primer set successfully amplified PC samples, shown in the amplification plot (Fig. S2). Melting-curve analyses of SARS-CoV-2 positive samples lead to a single peak, pointing out the generation of an abundant amplicon, rather than multiple unspecific- or spurious-priming products. Such abundant target specificity is also shown in Fig. 4b. Although LAMP reactions are known to produce multiple amplicons with randomized sizes, some amplicons could be more abundant, while rare ones could be masked, leading to a single peak at melting curve analyses. Indeed, it is known that PCR reactions follow the law of all-or-nothing. In other words, if a molecule is amplified in the first cycles, it will probably be logarithmically amplified, taking over the reaction. Yet, if a molecule is not amplified in such first cycles, it will likely not undergo such exponential amplification, not being eventually detected.

In addition, the assay’s cross-reactivity was tested against eight different respiratory viruses from different clinical samples, which included coronavirus 229 E, parainfluenza virus 3, parainfluenza virus 4, human metapneumovirus A+B, bocavirus, enterovirus and rhinovirus. The resulted colorimetric RT-LAMP tests showed color change only in SARS-CoV-2 samples, but not for the others. RT-LAMP products were then loaded in 2% agarose gel 40 min post-reaction. Only SARS-CoV-2 sample showed ladder-type patterns (Fig. S3). Both results indicate high specificity of this primer set against COVID-19 infection.

## Discussion

The colorimetric RT-LAMP technique’s simplicity allows for its use as a POC testing alternative for large-scale screening. Its diagnostic accuracy, however, must be comparable to the RT-qPCR gold-standard to consider such methodology reliable, avoiding false results and their undesired consequences. In the colorimetric detection, 94.1% of results were found to be consistent with RT-qPCR outcomes. However, three samples (S18, S30, and S32), among the tested specimens with the *S* gene primers set, contradicted the RT-qPCR results, and hence were considered inconclusive. Two of the samples (S30 and S32) were RT-qPCR positive without color development in colorimetric LAMP. In opposite, the other inconclusive sample (S18) was RT-qPCR negative, while color development was obvious upon LAMP reaction. Also, fluorometric LAMP led to consistent findings with colorimetric LAMP (Fig. S1). It is well known that the viral load has a great impact on colorimetric RT-LAMP and RT-qPCR sensitivity. The positive identification of S18 specimen by RT-LAMP, but not by RT-qPCR, points out the sensitivity of current assay. Consistency with RT-qPCR gold-standard results must be validated, to assure reliability of colorimetric RT-LAMP. The colorimetric RT-LAMP tests are reliable when testing samples with moderate to high viral loads, which usually corresponds to samples with Ct values lower than 40 in RT-qPCR. For example, over 700 clinical specimens were tested, via targeting both *N* and *RdRp* genes, showing color change in positive ones after 30 minutes reaction time at 65 °C with 97.5% sensitivity, for samples with high to moderate viral loads (Dao Thi et al. 2020). In that regard, most studies show agreement on colorimetric RT-LAMP-reaction time of 30 minutes (Amaral et al. 2021; Chow et al. 2020; Huang et al. 2020; Lalli et al. 2021; Promlek et al. 2022), with temperatures not exceeding 65 °C (Amaral et al. 2021; Aoki et al. 2021; Chow et al. 2020; Dao Thi et al. 2020; de Oliveira Coelho et al. 2021; Huang et al. 2020; Lalli et al. 2021; Luo et al. 2022; Nawattanapaiboon et al. 2021; Promlek et al. 2022), in moderate to high viral-load samples, to avoid false results. Authors discussed that reaction time over 30 minutes resulted in nonspecific amplification, and temperature settings above 65 °C gave false-positive results in their assays, especially in NTC (Aoki et al. 2021; Promlek et al. 2022). However, time and temperature settings vary depending on type of DNA-polymerase enzymes, primers, and samples in reaction mixtures. In this study, we used LavaLAMP RNA enzyme (Lucigen), which has an activation temperature of 68-74 °C. Based on melting temperature (T_m_) of primers, and after optimizing reaction conditions, 40 minutes at 70 °C was the optimum setting in our colorimetric RT-LAMP assay. In average, it takes around 20 min (Ct 40) to 37 min (Ct 74) for SARS-CoV-2 positive specimens to develop color. When compared with RT-qPCR results, colorimetric RT-LAMP results are consistent, except for S18, which was positive in colorimetric RT-LAMP, but not detected in RT-qPCR, highlighting the sensitivity of this technique. To better distinguish between positives and negatives, it was found that placing samples on ice immediately after RT-LAMP reaction enhanced color formation (de Oliveira Coelho et al. 2021). Overall, sensitivity of the colorimetric RT-LAMP reaction greatly depends on the assay’s LOD, sample quality, viral load, and time of sample collection from the disease onset (Promlek et al. 2022).

Furthermore, it is common to validate post-reaction amplifications by loading samples in agarose gel electrophoresis, and observe distinct band patterns in positive ones (Amaral et al. 2021; Aoki et al. 2021; Dao Thi et al. 2020; de Oliveira Coelho et al. 2021; Huang et al. 2020; Luo et al. 2022; Nawattanapaiboon et al. 2021). It was observed that intermediate results showed band patterns in AGE, indicating that amplification has occurred, regardless of color development. These samples may have low amplicon formation or a low pH reduction, being probably affected by the elution buffer used during the RNA extraction process (Aoki et al. 2021).

According to our results, the *N* and *S* gene primer sets showed the best analytical performance, and thus were chosen for further analyses. These primer sets were designed to target conserved (non-mutated) regions of the most recent VOC. So, this assay is expected to detect any SARS-CoV-2 variant, including the heavily mutated Omicron. Since most Omicron mutations are on the *S* gene, primer binding sites of such gene across VOC were screened (Fig. S4). Accordingly, 99.2% of sequences in SARS-CoV-2 Omicron (B.1.1.529) variant (among 136 nt in total primer length, one was found as variant/mutated) exhibited similarity of the ones for Delta (B.1.617.2) variant. Omicron’s specific point mutation (C/A) exists at LB binding site, which is five nucleotides away from the 3’-site of the primer. Although it is far from the critical 3’-end for the polymerase enzyme, and seems to be negligible for amplification, its potential effect on amplification of Omicron variant was examined. Indeed, this primer set successfully detected Omicron, which confirms that it targets a non-mutated region in the *S* gene. In addition, we confirmed the specificity of the primers against other respiratory viruses, and no cross-reactivity was observed.

Overall, the analytical performance of four primer sets targeting *N, S, RdRp* and *E* genes were elucidated. Among the tested genes, *N* and *S* had the highest analytical sensitivity when detected by both colorimetric and fluorometric RT-LAMP.

## Conclusions

Alternative COVID-19 diagnostic techniques to the RT-qPCR gold-standard are being broadly investigated, to address shortages in some areas. Colorimetric RT-LAMP has the potential to be used as a POC testing technique, showing a 94.6% sensitivity and 92.9% specificity comparable to RT-qPCR, with shorter reaction time and simpler implementation. Besides, it is cheaper, not requiring specialized equipment and trained personnel, and results can be interpreted by a simple color visualization. This LAMP test is expected to detect any SARS-CoV-2 variant, including the heavily mutated Omicron. That makes it an attractive candidate to be used in local emergency centers, and even for in-home testing.

## Data Availability

All data produced in the present work are contained in the manuscript

## Statements and Declarations

### Ethical approval

The study was approved by the Institutional Review Board (IRB) at Imam Abdulrahman bin Faisal University (IAU), with IRB number IRB-2020-13-406. All methods were carried out in accordance with current guidelines and regulations. De-identified samples left over after completion of diagnostic tests were used; hence, this study requires no consenting, as per institutional ethics committee regulations and informed consent.

## Acknowledgments

This study is partly funded by Institute for Research and Medical Consultations (IRMC; 2020-IRMC-S-3) and Deanship of Scientific Research (DSR) of Imam Abdulrahman bin Faisal University (IAU; COVID19-2020-026-IRMC).

