## Supplementary Files for "Colorimetric and fluorometric reverse-transcription loop-mediated isothermal amplification (RT-LAMP) assay for diagnosis of SARS-COV-2"

**Edited by Gabriel Dorado (17.7.22)**

**Galyah Alhamid^1^, Huseyin Tombuloglu^1^*, Dalal Motabagani^2^, Dana Motabagani^2^, Ali A. Rabaan^3,4,5^, Kubra Unver^6^, Gabriel Dorado^7^, Ebtesam Al-Suhaimi^8^, Turgay Unver^6*^**<--Update with data from the manuscript (fixed SPACES between names and numbers)

^1^ Department of Genetics Research, Institute for Research and Medical Consultations (IRMC), Imam Abdulrahman Bin Faisal University, 31441, Dammam, Saudi Arabia

^6^ Ficus Biotechnology, Ostim OSB Mah, 100. Yil Blv, No: 55, Yenimahalle, Ankara, Turkey<--Update with data from the manuscript (missing affiliations)

**Table S1.** Sequences of LAMP primers used in this study.

| ***Gene*** | **Sequence (5’ –> 3’)** |
| --- | --- |
| ***RdRp*** | RdRp-FIP: CCACCTGGTTTAACATATAGCTAATGAGTGTGCTCAAGTATTGAGTGAAA  RdRp-BIP: GTTTTTAACATTTGTCAAGCACCATCAGTAGATAAAAGTGCATTAACATT  RdRp-F3: GTAGCTTGTCACACCGTTTCTATAGATTAG  RdRp-B3: CATACTTATCGGCAATTTTGTT  RdRp-FL: CTAATGAGTGTGCTCAAGTATTGAGTGAAA  RdRp-BL: ACCATCAGTAGATAAAAGTGCATTAACATT |
| ***N*** | N-FIP: GCATTGTTAGCAGGATTGCGGGGTTGCAACTGAGGGAGCC  N-BIP: TCTACGCAGAAGGGAGCAGAGGGTTGCGACTACGTGATGAGG  N-F3: TGGTGCTAACAAAGACGGC  N-B3: TACTGCTGCCTGGAGTTGAA  N-FL: GTTGCAACTGAGGGAGCC  N-BL: GTTGCGACTACGTGATGAGG |
| ***E*** | E-FIP: GCGCAGTAAGGATGGCTAGTTACTTCTTTTTCTTGCTTTCGT  E-BIP: GTGCGTACTGCTGCAATATTGTTAAGAAGAATTCAGATTTTTAACACGAG  E-F3: AGGTACGTTAATAGTTAATAGCG  E-B3: ACCAGAAGATCAGGAACTCT  E-FL: TACTTCTTTTTCTTGCTTTCGT  E-BL: GAAGAATTCAGATTTTTAACACGAG |
| ***S**** | S-LB: TAAACCGTGCTTTAACTGGAATAGC  S-LF: ACTGATGTCTTGGTCATAGACACT  S-BIP: GTTGCAATATGGCAGTTTTTGTACATTGGGTGTTTTTGTCTTGTT  S-F3: TCTATTGCCATACCCACAA  S-B3: GGTGTTTTGTAAATTTGTTTGAC  S-FIP: CATTCAGTTGAATCACCACAAATGTGTGTTACCACAGAAATTCTACC |

F3: Forward outer; FIP: Forward inner; LF: Loop forward; B3: Backward outer; BIP: Backward inner; LB: Loop backward.<--Alphabetically sorted here?? *Yan et al. (2020).

To evaluate the reliability of the diagnostic test, Table S3 demonstrate the number of true positive (TP), true negative (TN), false positive (FP) and false negative (FN) values of all tested samples, in the colorimetric assay. The gold-standard RT-qPCR was used as reference (comparative method), to determine TP and TN results across all tested samples. Accordingly, the assays’ sensitivity, specificity, accuracy, positive predictive value (PPV) and negative predictive value (NPV) were determined based on the following formulas (Table S2):

**Table S2.** True positive (TP), false positive (FP), true negative (TN) and false negative (FN) terms utilized to evaluate the diagnostic accuracy of the RT-LAMP method.

| Comparative method | Test method  **+ -** | | Total |
| --- | --- | --- | --- |
| **+** | TP | FP | (TP + FP) |
| **–** | FN | TN | (FN + TN) |

Sensitivity = TP/(TP + FN)

Specificity = TN/(TN + FP)

Accuracy = (TP + TN)/(TP + TN + FP + FN)

PPA = TP/(TP + FP)

NPA = TN/(TN + FN)

**Table S3.** True positive (TP), false positive (FP), false negative (FN) and true negative (TN) results of 51 tested clinical-samples compared to the RT-PCR gold-standard.

| Comparative method | Test method  + - | | Total |
| --- | --- | --- | --- |
| + | 35 | 1 | 36 |
| – | 2 | 13 | 15 |

Based on Table S2 above, the following calculations were made:

Sensitivity = TP/(TP + FN) = [35/(35 + 2)]*100 = 94.6%
Specificity = TN/(TN + FP) = [13/(13 + 1)]*100 = 92.9%
Accuracy = (TP + TN)/(TP + TN + FP + FN) = [(35 + 13)/(35 + 13 + 1 +2)]*100 = 94.1%
PPV = TP/(TP + FP) = [35/(35 + 1)]*100 = 97.2%
NPV = TN/(TN + FN) = [13/(13 + 2)]*100 = 86.7%
Youden’s J index = sensitivity + specificity – 1 = 0.946 + 0.929 – 1 = 0.875

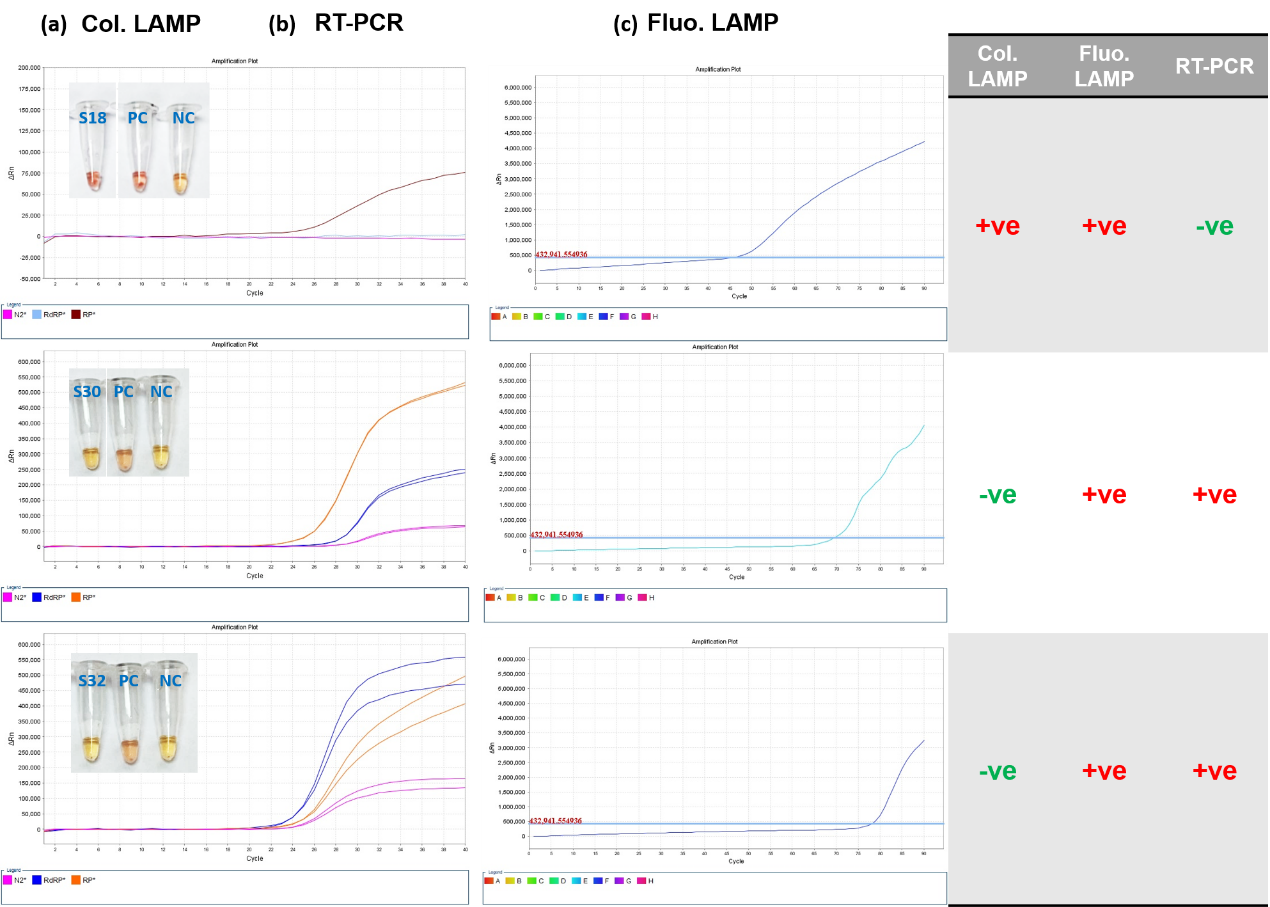

**Fig. S1.** Comparison of the three inconclusive results, based on (a) colorimetric LAMP, (b) RT-PCR and (c) fluorometric LAMP methods.

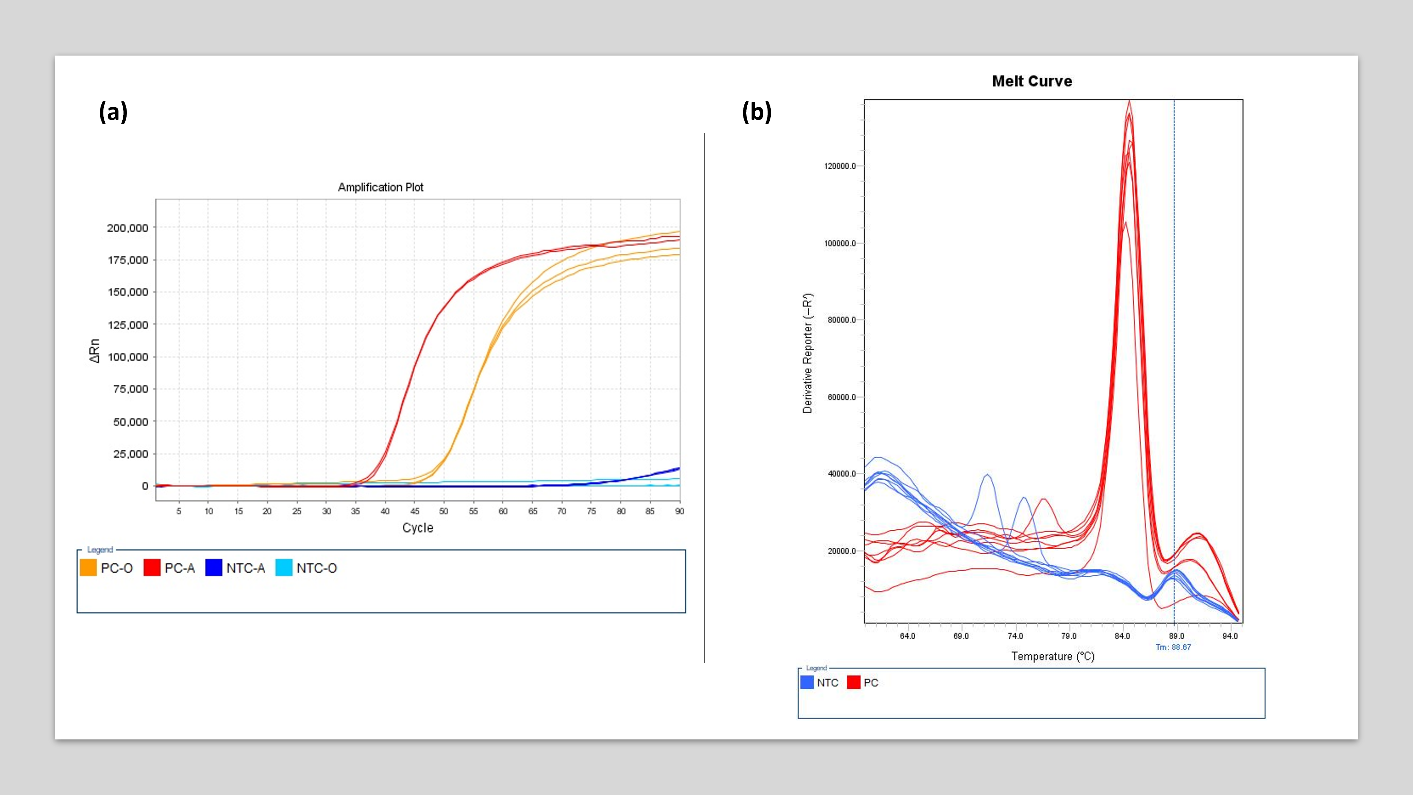

**Fig. S2. (a)** Fluorometric detection of *N* gene of SARS-CoV-2 positive samples. Three replicates of two different positive controls (PC-O, PC-A) and negative controls of the reactions (NTC-O and NTC-A). **(b)** Melting curve analyses of the same reactions.

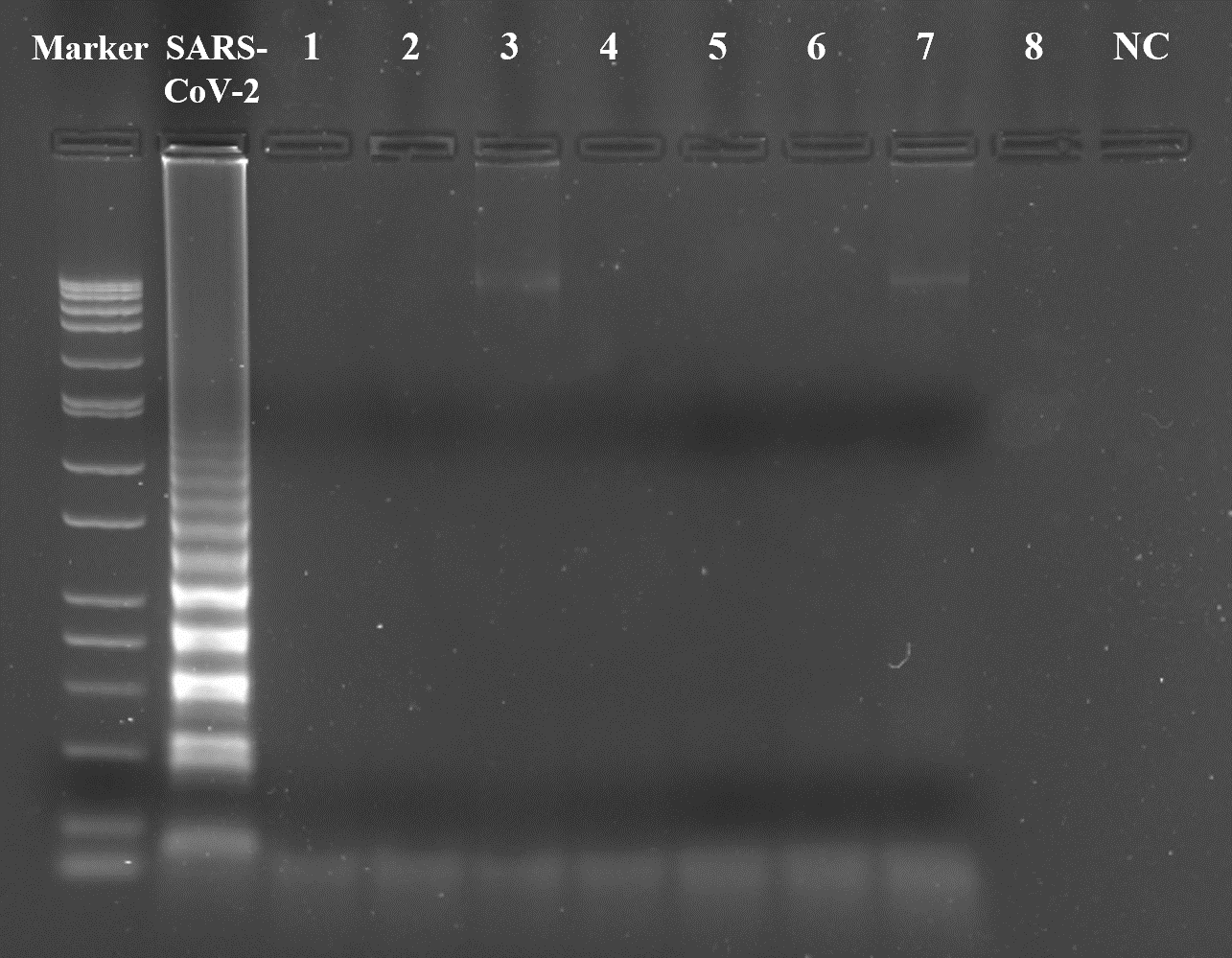

**Fig. S3.** Specificity of primers tested against clinical samples infected with SARS-CoV-2, along with other respiratory viruses (1-8). These include coronavirus 229 E, parainfluenza virus 3, parainfluenza virus 4, human metapneumovirus A+B, bocavirus, enterovirus and rhinovirus. Post-reaction gel electrophoresis with 2% agarose showed a ladder-type pattern in COVID-19-infected sample only, and nothing in samples with other respiratory viruses, indicating high specificity of the primers against SARS-CoV-2.

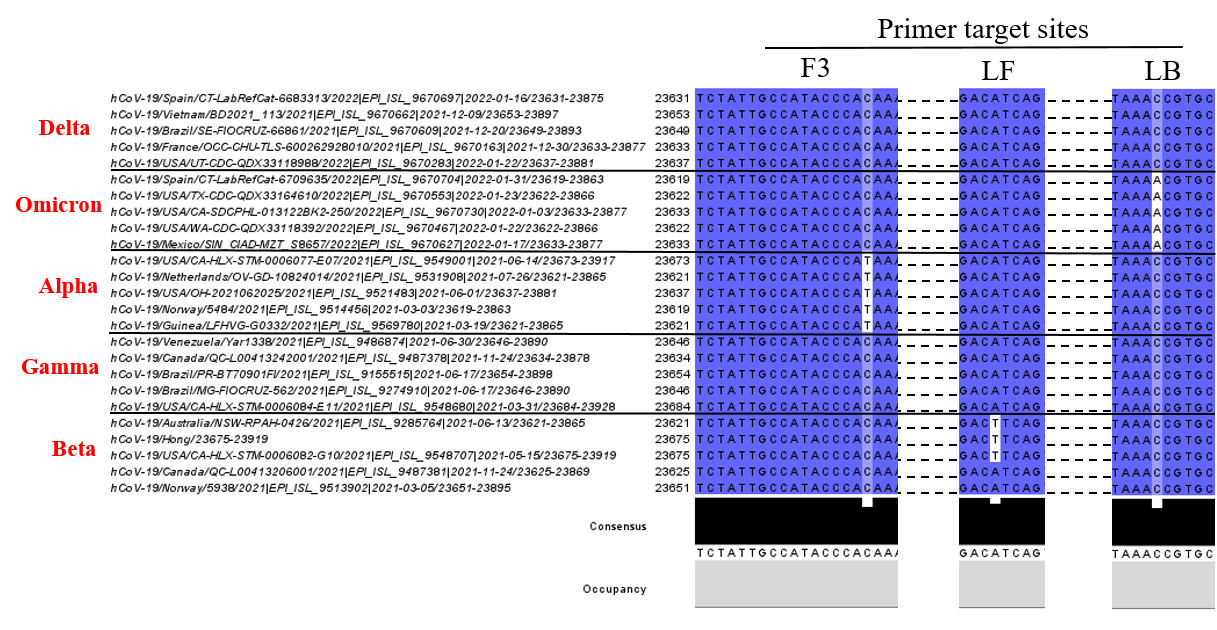

**Fig. S4.** Conservation of primer-binding sites of the *S* gene across VOC. Accordingly, there was 99.2% similarity of sequences (among 136 nt in total primer length, only one is different) from the *S* gene of SARS-CoV-2 Omicron (B.1.1.529) and Delta (B.1.617.2). A point mutation (C/A) exist at LB binding site.

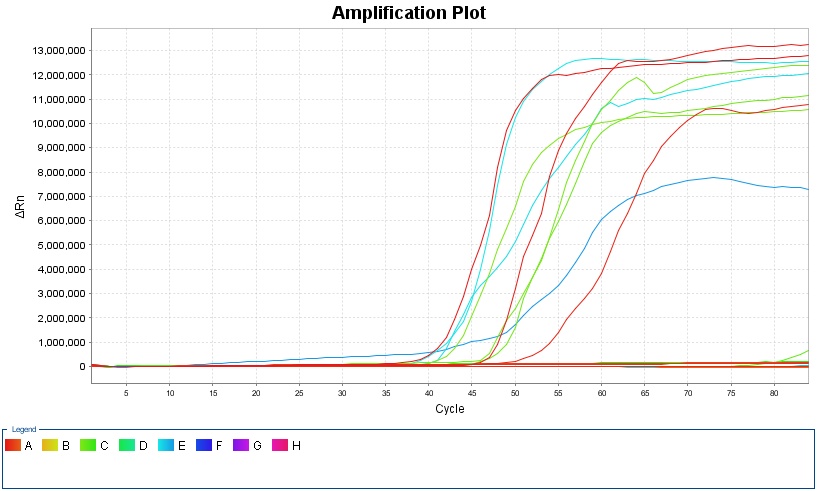

**Fig. S5.** Efficacy of S primers against the Omicron variant tested fluorometrically on nine clinical samples.	The assay successfully detected the samples infected with Omicron, which confirms the validity of the primers to identify SARS-CoV-2 variants.
